# Development of a comprehensive database of markers of ultra-processing to identify ultra-processed food in the UK according to the Nova classification

**DOI:** 10.64898/2026.09.24.26363859

**Authors:** Samuel J. Dicken, Dolly van Tulleken

## Abstract

Growing evidence links ultra-processed food intake according to the Nova classification with poor health, yet no comprehensive database of additives and food substances of non-culinary use that are markers of ultra-processing exists for the UK. Building the database involved seven steps: (1) obtaining all approved additives in the UK, (2) retrieving all of their technical functions, (3) coding each technical function as cosmetic (marker of ultra-processing) or non-cosmetic (not a marker of ultra-processing), (4) adding ‘flavour extracts’, ‘natural flavourings’, and ‘flavourings’, (5) excluding additives as ultra-processing markers if they served fortification purposes, or were a herb/spice, and (6) adding food substances of non-culinary use defined as markers of ultra-processing in the Nova classification. Lastly, (7) two UPF definitions were constructed based on the ambiguity/overlap of additive technical functions from previous approaches. The database is freely accessible for researchers to improve consistency in using the Nova classification in the UK.

## Background

Growing evidence links higher intakes of ultra-processed food (UPF) with poor health^1^. As most commonly conceptually defined by the Nova classification^2^, UPFs are branded, commercial formulations formulated using additives and food substances of non-culinary use (food substances never or rarely used in kitchens or culinary preparations). UPF are created to displace the other three Nova groups (unprocessed and minimally processed foods, processed culinary ingredients and processed foods) in people’s diets - and the culinary preparations associated with them - to maximise corporate profits^1^. UPF includes snacks, drinks, ready meals and other largely packaged products such as mass-produced breads, breakfast cereals, sweets and savoury snacks.

Operationally, the Nova classification identifies UPF by the presence of at least one marker of ultra-processing in the list of ingredients, including food substances generally not used in culinary preparations and classes of additives with cosmetic functions^2^.

Identifying UPF in research currently relies on using the list of examples of food substances and classes of additives in definition papers of the Nova classification^2,3^ plus best practice guidance to apply Nova to dietary assessment tools^4–6^.

In a recent UK Scientific Advisory Committee on Nutrition (SACN) statement on (ultra)-processed food, the Nova classification was the only food processing classifications considered to be potentially applicable to a UK population^7^. However, SACN highlighted concerns around interrater reliability to classify food and drink according to the Nova classification, and the need for assessment and refinement of future UK National Diet and Nutrition Survey (NDNS) methodology for monitoring UPF intake surveillance.

A key step in ensuring consistency and enhancing research rigour is to have a comprehensive list of country-specific markers of ultra-processing as defined by the Nova classification (given that specific additives or food substances of non-culinary use may or may not be approved or used in different countries). No such comprehensive list currently exists in the UK.

The aim of this research was to construct a freely available comprehensive database of all ultra-processing markers as defined by the Nova classification for application to food and drink in the UK.

## Methods

### Nova classification

The rationale and concept behind the Nova classification is discussed in detail elsewhere^1,2^. The distinction between the Nova classification and other food processing classifications in capturing the purpose behind processing (in contrast to the concept of the physical degree of processing), is detailed in Dicken, 2026^9^. In brief, the Nova classification uses a commercial determinants of health lens to help explain the transition towards less healthy diets in the past few decades and concomitant rise in diet-related non-communicable disease incidence, and the role of the increasingly globalised commercial food system. In order to capture these drivers in producing less healthy food and drink, the Nova classification uses product-level ingredient markers of additives and food substances of non-culinary use. The classification is not simply about their use *per se*, and attempts to capture the role of aggressive marketing, powerful branding, product placement, food price and ownership models under transnational corporations to help explain why and how traditional diets are being displaced by UPF and leading to poor health outcomes globally^1^. Further discussion on the validity and utility of Nova and the concept of ultra-processed foods for scientific research is outlined elsewhere^11^.

### Building the database

Development of the database was conducted according to the Nova classification and the 2026 Healthy Eating Research report ““Ultraprocessed Foods in the U.S.: Recommended Definitions and Policies”^12^, which published the methodology behind a comprehensive list of additives and food substances of non-culinary use that define markers of ultra-processing in the US^12^. Development of the database was conducted according to the approach outlined in Appendix C of the Healthy Eating Research report^12^ (Appendix C: Instructions on How to Develop a List of Current Ingredients and Additives Used for Identifying UPFs), which was applied to a UK context. The database building approach and specification of additive classes which define UPF in this study were confirmed with the authors of the Nova classification and Healthy Eating Research report authors.

Building the UK database of ultra-processing markers involved seven steps: (1) obtaining all approved additives in the UK, (2) retrieving all of their technical functions, (3) coding each technical function as cosmetic (marker of ultra-processing) or non-cosmetic (not a marker of ultra-processing) according to the Nova classification, (4) adding ‘flavour extracts’, ‘natural flavourings’, and ‘flavourings’ to the database, (5) excluding additives as markers of ultra-processing if they served fortification purposes, or were a herb or spice as per guidance^12^, and (6) adding food substances of non-culinary use defined as markers of ultra-processing in the Nova classification^2^ and Healthy Eating Research report^12^. Lastly, (7) two definitions of UPF were constructed based on the ambiguity/overlap of additive technical functions based on previous approaches: (a) additive present in the product is a marker of ultra-processing if it has any cosmetic technical function (US definition)^2,12^, (b) additive present in product is a marker of ultra-processing only if the specified function on the ingredients list is a cosmetic technical function according to the Nova classification (UK definition)^12^.

For step 1, a complete list of additives approved for use in the UK were obtained from the Food Standards Agency (FSA) government website^13^, and exported into Excel (Microsoft). Additives are substances not normally consumed as a food by themselves and not normally used as typical ingredients in foods. They are added to foods to provide a specific technological purpose, such as extending shelf life, preventing microorganism profilferation, or improving or enhancing the sensory properties of food^14^.

For step 2, all technical functions for the list of FSA-approved additives were retrieved. Additives can have more than one technical function, as defined by the Food and Agriculture Organization of the United Nations / World Health Organisation (FAO/WHO) Codex Alimentarius^15^. The Nova classification uses these technical functions to define markers of ultra-processing. However, the FSA only lists only one function for a number of approved additives, and groups most additives together into a single list with unclear specification of their function^13^. To ensure a complete list of all technical functions of approved additives in the UK, the WHO/FAO Codex Alimentarius database of all additives and their functions was accessed^15^. This database includes additives that are not approved for use in the UK, therefore only the additives that are approved in the UK were checked. All technical functions were retrieved and assigned to the respective UK FSA-approved additive in Excel. Additionally, alternative names for each additive were also retrieved to maximise generalisability and capture the possible range of wording used in ingredients lists (e.g. E132: Indigotine, and Indigo Carmine).

For step 3, each technical function in the Excel database for each additive approved for use in the UK was coded as cosmetic (i.e. a marker of ultra-processing) or non-cosmetic (i.e. not a marker of ultra-processing) according to the Nova classification. Out of the 28 technical classes of additives in WHO/FAO Codex Alimentarius (27 plus flavours, which are evaluated by the Joint FAO/WHO Expert Committee on Food Additives (JECFA) and are not included in the Codex International Numbering System for Food Additives (INS)), 13 are considered to be ‘cosmetic additives’ to define UPF, which capture the purpose of ultra-processing. As outlined in Monteiro et al 2019, the function of these cosmetic additives’ is proposed to make the final product palatable or often hyper-palatable^2^, and impart sensory properties that would not be present in the final product without the additive. These 13 technical classes are: anti-foaming agents, bulking agents, carbonating agents, colours, emulsifiers, emulsifying salts, flavours, flavour enhancers, foaming agents, gelling agents, glazing agents, sweeteners and thickeners.

The other WHO/FAO Codex Alimentarius technical classes are not considered to be ‘cosmetic additives’ and are not markers of ultra-processing. Their presence alone does not mark a product as UPF. These technical classes are: acidity regulators, anti-caking agents, antioxidants, bleaching agents, carriers and carrier solvents, colour-retention agents and colour stabilisers, firming agents, flour treatment agents, humectants, packaging gases, preservatives, propellants, raising agents, sequestrants, and stabilisers.

The 27 technical classes of additives (and flavours as evaluated by JEFCA) in WHO/FAO Codex Alimentarius are listed and coded into cosmetic (i.e. a marker of ultra-processing) or non-cosmetic (i.e. not a marker of ultra-processing) in Table 1.

**Table 1.**
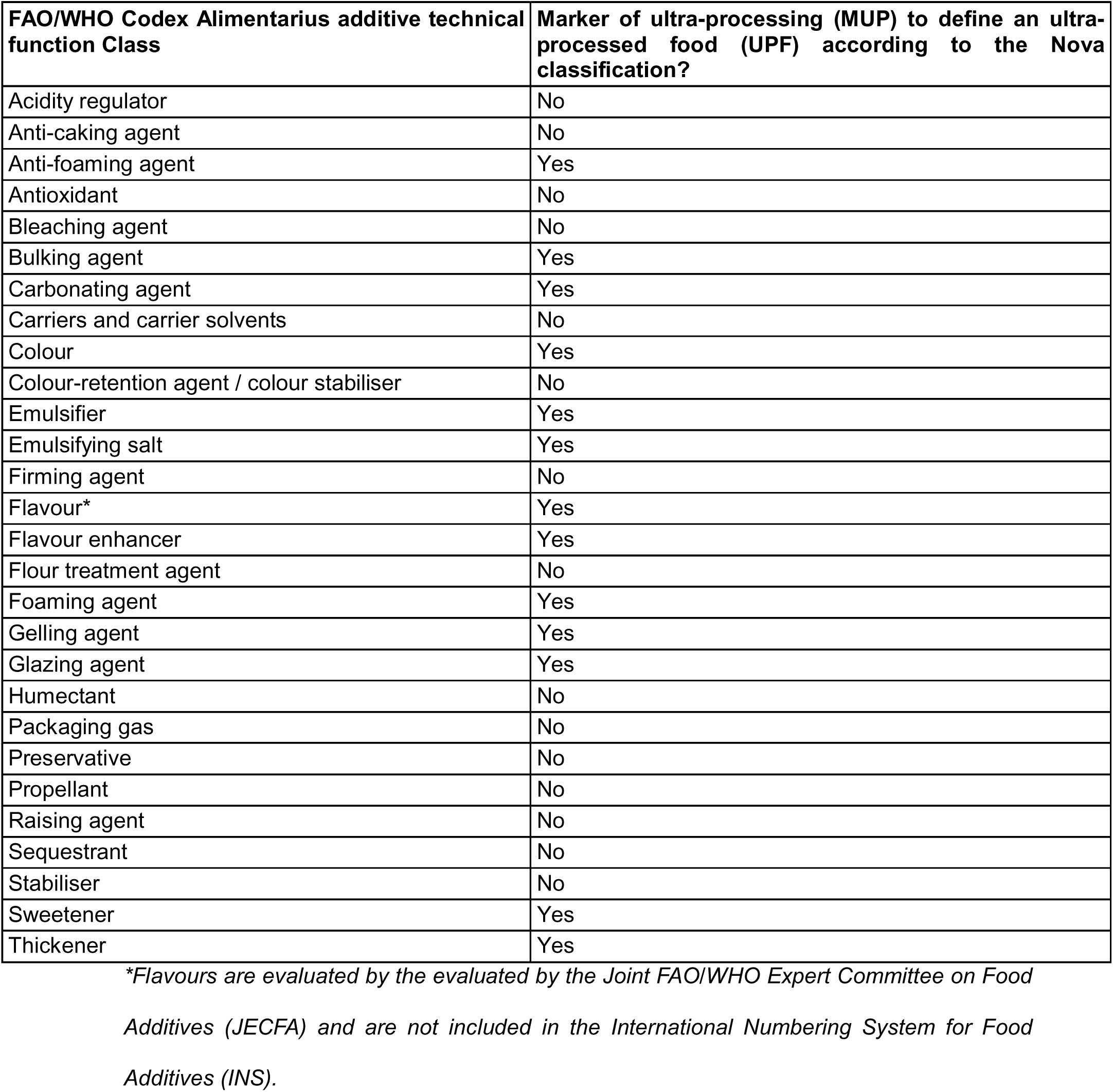
FAO/WHO Codex Alimentarius classes of additives with cosmetic functions that the Nova classification considers to be markers of food ultra-processing (MUP)

Where necessary, European Food Safety Authority (EFSA) Scientific Opinions were consulted if the additive was not explicitly listed in WHO/FAO Codex Alimentarius (e.g. E472f: Mixed acetic and tartaric acid esters of mono-and diglycerides of fatty acids), as well as comparing technical functions of near-identical additives in WHO/FAO Codex Alimentarius (e.g. E472e: Mono- and diacetyl tartaric acid esters of mono-and diglycerides of fatty acids).

For step 4, ‘flavour extracts’, ‘natural flavourings’, and ‘flavourings’ were added to the database. Flavours are not included in WHO/FAO Codex Alimentarius. In the UK, the technical function of flavourings must be specified in the ingredients list (e.g. ‘flavouring’), but the specific flavouring name does not have to be specified and is rarely provided. Therefore, as all flavouring substances are defined as markers of ultra-processing according to the Nova classification, the generic terms ‘flavour extract’, ‘flavouring’ and ‘natural flavouring’, were added to the database as rows, as similarly conducted in the Healthy Eating Research report^12^. There are 2501 approved flavourings in the UK^16^. The full list was added to the Excel database in a separate sheet for completeness.

For step 5, additives were excluded as markers of ultra-processing if they served micronutrient fortification purposes, or were a herb or spice, as per guidance^12^. As outlined in the Healthy Eating Research report^12^, some additives are excluded from being a marker of ultra-processing based on their wider function or use. This includes additives that serve micronutrient fortification purposes, and additives that are household herbs and/or spices. These were identified using UK specific documents. Vitamins and minerals were identified using the UK government ‘Great Britain register on the addition of vitamins and minerals and of certain other substances to foods’^17^. For herbs and spices, there is no UK government document. However, FSA documentation on herbs and spices^18^ links to the Seasoning and Spice Association (SSA) website, which contains a list of culinary herbs and spices^19^. This list was used to identify and exclude any herbs and spices as a marker of ultra-processing. Sodium chloride and yeast and yeast-derived additives are also excluded as markers of ultra-processing according to the Healthy Eating Research report. There were no sodium chloride and yeast and yeast-derived additives in the FSA list of approved additives (i.e. none in the database to exclude).

For step 6, food substances of non-culinary use were added to the database in Excel. Food substances of non-culinary use are sources of energy and nutrients that are never or rarely used in domestic kitchens^20^. They are all considered to be markers of ultra-processing, and include isolated extracts and include sugars, starches, fats and proteins. Some food substances of non-culinary use may also function as additives. Food substances of non-culinary use were retrieved from the definition papers of the Nova classification^2,3^ and Healthy Eating Research report^12^. Additional food substances of non-culinary use were identified from a published paper reporting such substances in the UK (but which does not report a full list of additives that are markers of ultra-processing)^10^.

For step 7, two definitions of UPF were constructed based on the different reporting of additive functions on ingredients lists in the UK compared with other countries such as the US. In the US, the primary technical function of an additive does not have to be specified by the manufacturer on the product ingredients list. Therefore, a liberal definition assumes that the presence of an additive on a product ingredients list that has one or more possible functions that can be cosmetic or non-cosmetic is always a marker of ultra-processing. For example, sodium alginate (E401) can act as an emulsifier (a marker of ultra-processing) and a humectant (not a marker of ultra-processing). In this definition, the presence of sodium alginate (i.e. an additive that can function as an emulsifier) would always define the product as UPF.

In the UK, the primary technical function of the additive must be specified by the manufacturer on the product ingredients list. Therefore, a conservative estimate can be made to only define the presence of an additive as a marker of ultra-processing if the specified technical function is a cosmetic function according to the Nova classification (Table 1)^10^. In this case, sodium alginate (E401) would only define the product as UPF if it was reported on the ingredients list as an emulsifier (a marker of ultra-processing), but not if it was reported on the ingredients list as a humectant (not a marker of ultra-processing).

### Analysis

Upon completion of the database, descriptive analyses were performed, including reporting: (1) the number of additives and food substances of non-culinary use in the database, (2) for additives (including flavours): (2i) the average number and range of technical functions, (2ii) the average number and range of cosmetic and non-cosmetic technical functions, and (2iii) the number of additives with both cosmetic and non-cosmetic technical functions, (3) the number of markers of ultra-processing according to the US definition, (4) the number of definite markers of ultra-processing according to the UK definition, (5i) the number of possible markers of ultra-processing according to the UK definition (depending on the specified technical function), of which, (5ii) the types of additives that are possible markers of ultra-processing based on the FSA reported technical function, and, (5iii) the average number and range of cosmetic and non-cosmetic technical functions of possible markers of ultra-processing. Lastly, (6) the number of additives that are definitively not a marker of ultra-processing in any context.

The database was built in Microsoft Excel (Version 16.110.2 (26062818)).

## Results

In total, there were 369 items in the database. Of which, 335 were additives (including 3 generic flavour terms), and 34 were food substances of non-culinary use (Table 2). The entire database is provided in the Supplementary Materials. Out of 369 total items, 42 additives were excluded as markers of ultra-processing (one herb and/or spice, and 41 micronutrients). Of the remaining 327 items (293 additives including 3 generic flavour terms) after exclusions, 99 additives were definitively not a marker of ultra-processing in either US or UK definitions (i.e. all technical functions were non-cosmetic)

**Table 2:** Overview of the database of markers of ultra-processing for the UK by definition.

|  | Total number of | Additives (including | Food substances of |
| --- | --- | --- | --- |

|  | items | generic flavour terms) | non-culinary use |
| --- | --- | --- | --- |
| **Total database** | 369 | 335 | 34 |
| Excluded as a marker of ultra-processing (41 micronutrients, 1 herb/spice) | 42 | 42 | 0 |
| Not a marker of ultra-processing (US and UK definitions) (additive has no cosmetic technical functions) | 99 | 99 | 0 |
| **US definition (N = 228)** |  |  |  |
| Marker of ultra-processing | 228 | 194 | 34 |
| **UK definition (N = 228)** |  |  |  |
| Marker of ultra-processing | 125 | 91 | 34 |
| Possible marker of ultra-processing (additive has both cosmetic and non-cosmetic technical functions) | 103 | 103 | 0 |

Of the remaining 228 items, all were markers of ultra-processing according to the US definition (i.e. the presence of an additive on a product ingredients list that has one or more cosmetic functions is always a marker of ultra-processing), including 194 additives and 3 generic flavour terms.

Out of the same 228 items, 125 items were definite markers of ultra-processing according to the UK definition (i.e. the additive present in product is only a marker of ultra-processing if the specified technical function on the ingredients list is a cosmetic function according to the Nova classification, and therefore only a definite marker if all its technical functions are cosmetic), including 88 additives and 3 generic flavour terms.

The remaining 103 additives out of 228 items were all possible markers of ultra-processing according to the UK definition, depending on the specified technical function on the product ingredients list (i.e. contains both cosmetic and non-cosmetic technical functions). According to the FSA primary listing, these were sweeteners (N = 8), emulsifiers, stabilisers and gelling agents (N = 55), and other: acidity regulators, anti-caking agents, anti-foaming agents, bulking agents, carriers and carrier solvents, emulsifying salts, firming agents, flavour enhancers, flour treatment agents, foaming agents, glazing agents, humectants, modified starches, packaging gases, propellants, raising agents and sequestrants (N = 40).

Table 3 reports the cosmetic and non-cosmetic additive technical functions according to the Nova classification by US and UK definitions.

**Table 3.**
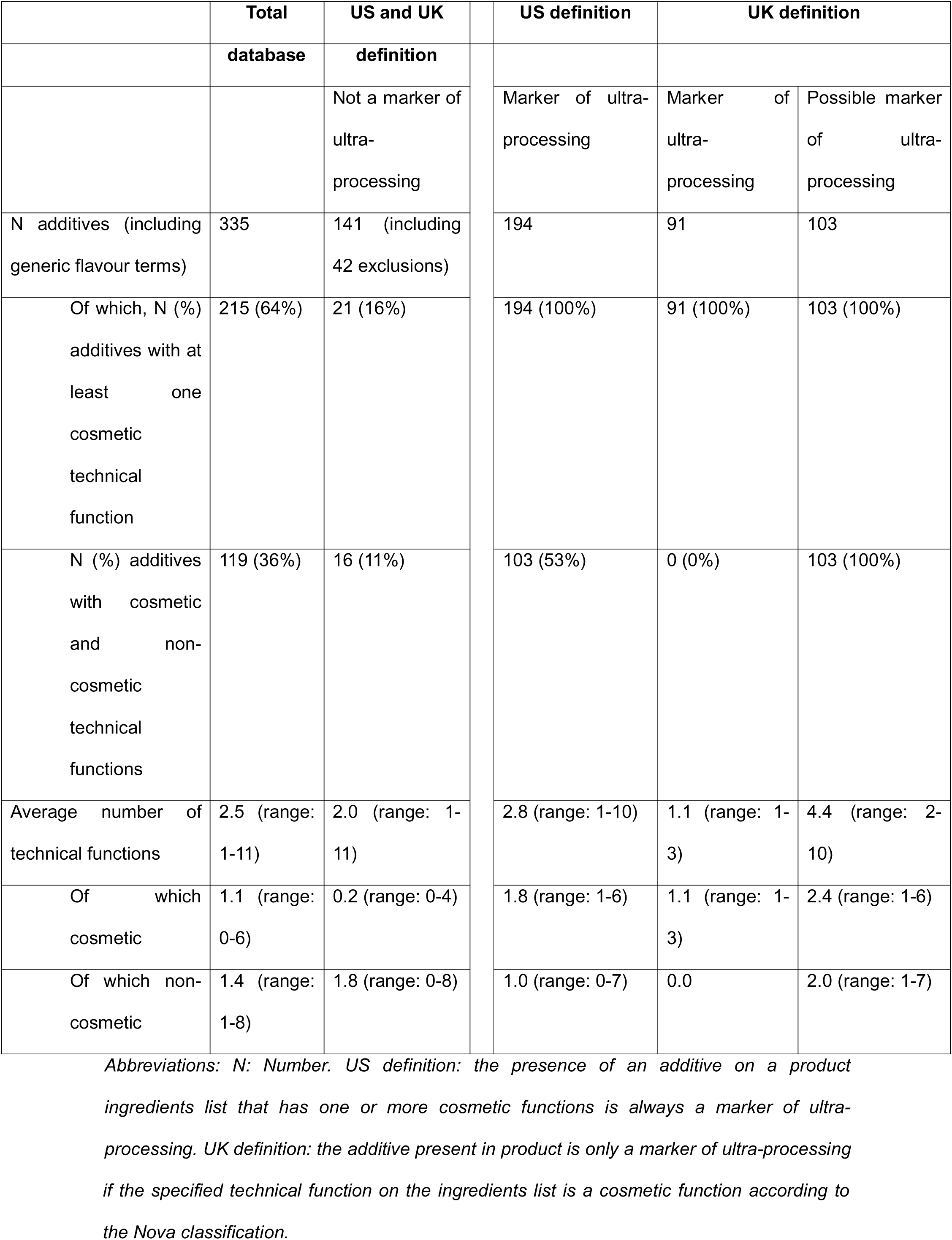
Summary of cosmetic and non-cosmetic additive technical functions according to the Nova classification by UK and US definitions.

Across 335 additives including 3 generic flavour terms, there was an average of 2.5 technical functions (range: 1-11). Of which, there were 1.4 non-cosmetic functions (range: 1-8), and 1.1 cosmetic functions (range: 0-6). There were 215 additives with at least one cosmetic technical function, of which, 119 additives had both cosmetic and non-cosmetic technical functions.

For the 141 additives that were not markers of ultra-processing according to the US and UK definitions, these additives had an average of 2.0 technical functions (range: 1-11): 1.8 non-cosmetic technical functions (range: 0-8), and 0.2 cosmetic technical functions (range: 0-4).

For the 194 additives including 3 generic flavour terms that were markers of ultra-processing according to the US definition, there was an average of 2.8 technical functions (range: 1-10). Of which, there were 1.0 non-cosmetic functions (range: 0-7), and 1.8 cosmetic functions (range: 1-6). Of the 194 additives, 103 (53%) had both cosmetic and non-cosmetic technical functions.

For the 91 additives including 3 generic flavour terms that were markers of ultra-processing according to the UK definition, there was an average of 1.1 technical functions (range: 1-3), all of which were cosmetic functions.

For the 103 additives that were possible markers of ultra-processing according to the UK definition, these additives had an average of 4.4 technical functions (range: 2-10): with 2.0 non-cosmetic technical functions (range: 1-7), and 2.4 cosmetic technical functions (range: 1-6).

## Discussion

This study provides the first freely available comprehensive database of additives and food substances of non-culinary use that are markers of ultra-processing according to the Nova classification in the UK. This database was constructed following a recently published database approach in the US according to the Nova classification and was confirmed through discussion with the creators of the Nova classification definition.

There is potential that some researchers when trying to apply the Nova classification have used a different set of markers of ultra-processing. There has also been debate around defining UPF^8^, including confusion between different processing classification systems (discussed in detail elsewhere^9^). This freely accessible database therefore serves as a useful tool for researchers, policymakers and other stakeholders, with the aim of improving consistency and enhancing rigour in coding food and drink into the Nova classification in the UK.

A given additive can serve multiple technical functions in a product. Due to country-specific reporting of the primary role of additives in the UK versus US, different approaches to defining UPF based on the specified function of the additive, or presence of the additive *per se,* are possible. In the UK, the additive technical function must be specified on the product. As outlined by WHO/FAO, it is the responsibility of the food manufacturer, not regulatory authority, to declare which functional class is most appropriate in the ingredients list. In the UK food supply, a recent paper indicated that 90% of UPF can be identified with three technical classes of additive: flavour (58% of all UPF), emulsifiers (36% of all UPF) and colours (27% of all UPF)^10^. However, this study used the US definition to define UPF, and thus it is unclear how identification with the UK definition would differ in capturing UPF. Future research can test the relative impact in defining UPF between both US and UK definitions.

In a recent UK SACN statement on (ultra)-processed food, out of eight food processing classifications, the Nova classification was the only system considered to be potentially applicable to a UK population^7^. As such, the development of this open-access database of markers of ultra-processing provides a key resource to facilitate future research in a UK context that can reliably be applied to estimate consumption of UPF. This can be used to address current limitations, such as investigating inter-rater coding reliability, assessment and refinement of future NDNS methodology for monitoring UPF intake surveillance, assessment of the robustness of the Nova classification, and development and validation of Nova-specific dietary assessment tools and validation of previous estimates of UPF intake.

The markers of ultra-processing are not in themselves proposed to be the primary causal mechanisms linking UPF intake and health outcomes. This database facilitates future work to examine the utility of these markers in capturing broader mechanisms.

Strengths of this study include being the first freely available comprehensive database of additives and food substances of non-culinary use that define markers of ultra-processing according to the Nova classification in the UK. The approach was aligned with a recent US database and confirmed with the lead author of the Nova classification. This ensures the database is precisely aligned with the Nova classification, providing researchers, policymakers and other stakeholders with a reliable resource for research related to the Nova classification. The database also includes the E number of additives and multiple alternative names and synonyms to maximise applicability and use, with an accompanying set of notes on how to navigate and use the database. The database contains a full breakdown of data sources, cosmetic/non-cosmetic additive technical functions of all FSA-approved additives, and reasons for exclusions. Given the adapting nature of the food industry, product reformulation and food regulation, this database is designed to be easily adaptable to accommodate changes, such as approvals of any new additives, withdrawals, specification of new technical functions, or changes to ingredients used in the UK. The detail provided in the database also ensures that it can be used for a range of settings and purposes, such as building new dietary assessment tools or coding existing dietary assessment tools, scanning product ingredient lists in retail settings, or combining with nutrient databases.

Limitations include that the database may not be generalisable to other countries, particularly those outside of the EU. Whilst the list of approved additives in the UK is largely based on EFSA recommendations, the exact list and range of food substances of non-culinary use may differ. However, given the close similarities between UK and EU additive approvals, the list will have reasonable utility and can be adapted to other countries in Europe.

## Conclusions

This study provides the first freely available comprehensive database of additives and food substances of non-culinary use that are markers of ultra-processing according to the Nova classification in the UK. The database is freely accessible for researchers to improve consistency in coding food and drink when applying the Nova classification in the UK.

## Availability of data and materials

The complete dataset supporting the conclusions of this article are available via UK Data Service (https://reshare.ukdataservice.ac.uk/858706/). The database includes a summary sheet and user guide how to use the database and data variables. The database is for research and educational purposes only, any commercial use requires approval and a license from the Authors. Please cite this paper when used.

## Code availability

Not applicable. No code was used or created in this analysis.

## List of abbreviations

UPF: Ultra-processed food
EFSA: European Food Safety Authority
FAO: Food and Agriculture Organization of the United Nations
JEFCA: Joint FAO/WHO Expert Committee on Food Additives
INS: International Numbering System for Food Additives
N: Number
NDNS: National Diet and Nutrition Survey
SACN: Scientific Advisory Committee on Nutrition
WHO: World Health Organisation

## Author contributions

SD: conceptualisation, data curation, data analysis, writing – first draft. DvT: conceptualisation, writing – review. All authors have read and agree to the final manuscript.

## Ethics approval and consent

Not applicable. This study did not involve any human participants or animals, no ethical approval or consent statement is required.

## Consent for publication

Not applicable.

## Acknowledgements

We thank Carlos Monteiro, Lindsey Smith-Taillie and Bridget Hollingsworth for their constructive comments on designing the database and for confirming the approach alignment with the Nova classification, and Carlos Monteiro for also providing constructive comments on the manuscript.

The authors would also like to thank the Food, Farming and Countryside Commission (FFCC) - conceptualisation of this database and manuscript arose whilst conducting work funded by the FFCC, which was then subsequently constructed by the authors independent of funding.

